# Partial unlock for COVID-19-like epidemics can save 1-3 million lives worldwide

**DOI:** 10.1101/2020.04.13.20064139

**Authors:** Robert L. Shuler, Theodore Koukouvitis, Dyske Suematsu

## Abstract

**Background:** A large percentage of deaths in an epidemic or pandemic can be due to overshoot of population (herd) immunity, either from the initial peak or from planned or unplanned exit from lockdown or social distancing conditions.

**Objectives:** We study partial unlock or reopening interaction with seasonal effects in a managed epidemic to quantify overshoot effects on small and large unlock steps and discover robust strategies for reducing overshoot.

**Methods:** We simulate partial unlock of social distancing for epidemics over a range of replication factor, immunity duration and seasonality factor for strategies targeting immunity thresholds using overshoot optimization.

**Results:** Seasonality change must be taken into account as one of the steps in an easing sequence, and a two step unlock, including seasonal effects, minimizes overshoot and deaths. It may cause undershoot, which causes rebounds and assists survival of the pathogen.

**Conclusions:** Partial easing levels, even low levels for economic relief while waiting on a vaccine, have population immunity thresholds based on the reduced replication rates and may experience overshoot as well. We further find a two step strategy remains highly sensitive to variations in case ratio, replication factor, seasonality and timing. We demonstrate a three or more step strategy is more robust, and conclude that the best possible approach minimizes deaths under a range of likely actual conditions which include public response.

## 1. Introduction

Epidemiological models produce fear-inducing scenarios that often persuade people to take dramatic mitigating steps, resulting in falsification of the model and undermining public confidence in experts. Hurricanes, after all, do not change their track because of a prediction. Attempts to model the actions of people still face the same dilemma. The situation is further mystified by a hidden agent meddling in the process of managing the epidemic, seasonality. For a new pathogen, seasonality may be unknown. By the time it is discovered, it has been affecting replication rate for some time.

The general goals of global efforts against a pandemic are threefold: (1) reduce the number of infected individuals and therefore deaths, (2) avoid overtaxing the healthcare system (which would restrict all services not just COVID-19), and (3) reduce the social and economic impact of the pandemic [1].

In the case of COVID-19 ballooning case rates have been brought down, but it is generally conceded that the effort is failing on the third count, social and economic impact. As regards the second goal, the world healthcare system for elective procedures is already shut down.

In regard to the first goal, the eventual death toll may not be reduced by mitigating action unless it can be sustained until a vaccine is available. One promising vaccine developer says it can produce 100 million doses a year and maybe one billion through alliances (https://www.wsj.com/articles/modernas-vaccine-hope-11589835889). It would take half a dozen such efforts to effectively vaccinate the world population within a year of starting, or at least 18 months from initial lockdown. The UN already says 125 million additional people are at risk of starting due to COVID-19 (https://www.bbc.com/news/world-52373888).

There are three causes of death due to lockdown which suppresses human survival activity: (a) mortality among cases that occur ahead of the development of a vaccine, if a vaccine is developed, but the associated total cases slow further spread; (b) mortality among overshoot cases, which contributes little; (c) deaths due to social unrest or economic conditions, which we do not quantify in this paper, but which could be large. By some accounts the Arab Spring including the Libyan, Yemen and Syrian civil wars, as well as civil war in Ukraine and massive migration into Europe, can be traced to economic disturbances somewhat more subtle than 18 months (potentially) of global shutdown [2].

In this paper we consider all strategies except remaining fully locked down for 18 months. In particular we examine whether overshoot prevention is necessary for small unlock steps of 10% or 20%, and whether the additional cases from such small steps are small enough that continued restraint is actually merited. And we study what kind of unlock schedule results in robust performance across the various unknowns in the early stages of an epidemic.

## 2. Approach

We use a model of epidemic spread with forcing functions for seasonality and social factors (lockdown). The model self-calibrates social factors from input data. We then consider various strategies looking for unnecessary deaths and developing methods to prevent them.

The methods our approach might reveal would be employed after containment is an opportunity past and a vaccine is a prospect too far in the future to avoid economic catastrophe. Opinions differ as to the effect of severe and prolonged recession on mortality and health. For example, there are fewer motorway deaths due to less driving [3]. During the COVID-19 pandemic there may well be fewer deaths due to pollution. On the other hand, the 2008 financial crisis resulted over the next few years in at least 260,000 additional cancer deaths [4]. Economic losses from pandemics, even without a long term global shutdown, have been estimated at the low end of but within the range of impacts from climate change [5]. These historical analyses are likely to vastly underestimate the impact from the economic and social disruption of COVID-19.

That leaves the approach of curve flattening, which can have one or both of two objectives as they lie on a continuous spectrum:

Keep the number of cases extremely low (and in consequence the economy completely shut down) until someone develops a therapy that prevents the disease or dramatically lowers mortality, or until the disease disappears on its own (unlikely if no herd immunity is building).

Keep the number of cases moderately low while herd immunity builds more slowly, but the medical system remains operational, the economy is not fully shut down, and the length of shutdown is minimized.

At relatively low case levels for COVID-19 in March 2020, fear-based shutdown was evidenced by traffic reductions in cities that had no shutdown nearly equal to that of cities which were locked down (See https://www.bbc.com/news/world-52103747 and scroll to “Travel declines even without official lockdowns”). Figure 1 shows the early replication rate history of COVID-19, with R_0_ defined as (new cases) / [(active cases) *(spreading days)].

**Figure 1.**
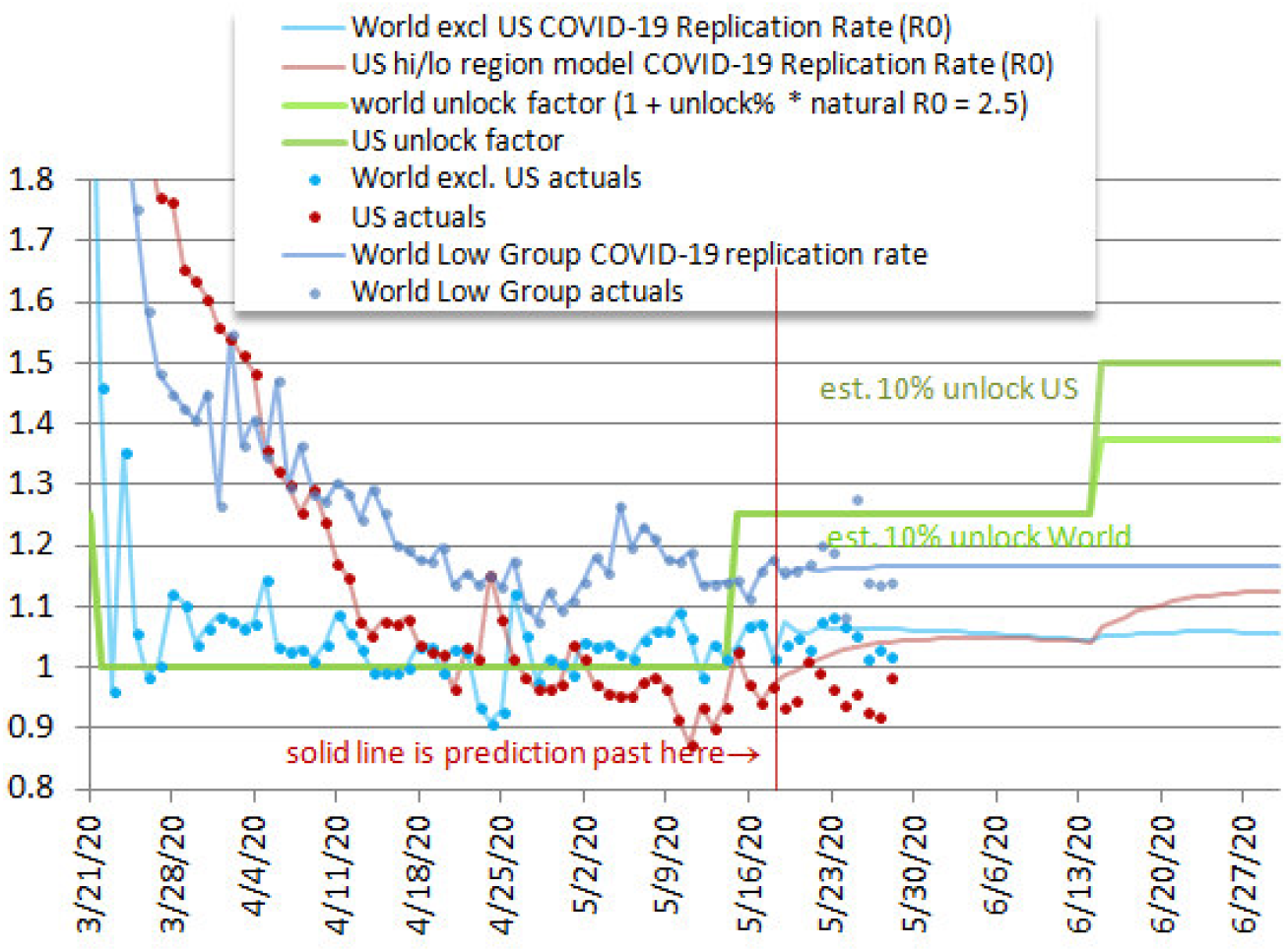
Empirical replication rate R_0_ for US, World-excluding-US, and World-Low-Group countries during phase-in of lockdown and social distancing – 4-day moving average. World-Low-Group had less than 100 deaths/million population on 5/18/2020, China excluded (Saint Lucia India Uzbekistan Ivory Coast Comoros Tunisia Congo Palestine Trinidad and Tobago Indonesia Sierra Leone Sudan Haiti Jordan Macao New Caledonia South Sudan Sri Lanka Liberia Mali Zambia Togo Belize Thailand Mauritania Mongolia Nicaragua Chad Niger Burkina Faso Nigeria Venezuela Bhutan Rwanda DRC Kenya Nepal Fiji Taiwan Suriname Madagascar Timor-Leste Botswana Benin Libya Eritrea Gambia Western Sahara Tanzania Namibia Yemen Cambodia Mozambique Ethiopia Myanmar Uganda Malawi Syria Zimbabwe Burundi Vietnam Laos Angola Papua New Guinea Lesotho).

While lockdowns began to be enforced outside China from late January, they were initially selective and directed at international travel. By mid-March they were widespread. We would expect a few days delay before a reduction in R_0_ would appear in the new cases data (taken from the CDC, from https://www.worldometers.info/coronavirus/, and historical data from https://github.com/nytimes/covid-19-data). Prior to March 21 the US cases were doubling every two days for about a week. This quickly declined following March 21, so that date was chosen as the start date for our tracking and modeling.

The chart suggests case data lags from changes in lockdown of a week or slightly less. Lockdowns were mostly in place by the end of March. Effectiveness of lockdown appears to have reached a maximum by mid-April. We believe the downturn in US R_0_ in May likely represents seasonal effects which are predicted to be as much as 40% in New York and 20% in Florida [6]. World R_0_ reflects mixed seasonality and approaches.

It is possible there would be little response if governments eased lockdown, unless they also declared the environment safe, which isn’t true any time soon. However, as economic distress builds, coupling easing with the ability to work would likely be a powerful motivator. If the initial drop on the left is the response to government recommendations as we speculate, then it may be indicative of the control flexibility over the lockdown replication rate.

The authors of this paper believe that such measures should be voluntary and regional. Those healthy and at low risk and in economic need are likely to be willing to expose themselves to the environment, especially if governments maintain the integrity of the healthcare system and people are not dying from neglect.

At this writing, many countries are implementing or considering partial unlock. This might result in a gradual approach to the minimum cases or a dramatic overshoot and unnecessary deaths. The intent of such moves is to slowly return to normal. Rebounds approach the herd immunity threshold too fast and cause overshoot [7, 8]. Some investigators refer to rebounds as a second outbreak or second wave, and specifically identify that it is likely to be uncontrolled and cause significant overshoot [9].

### 2.1. Model parameters

The model is in a spreadsheet to be uploaded with this paper, and also available at http://shulerresearch.org/covid19.htm. The particular parameters for COVID-19 are explained below. Some of the parameters are varied over a range making the results applicable to other similar epidemics. If the R_0_ is more than about 5 then it may peak before unprepared governments can respond. Otherwise our general conclusions should apply to some degree. All parameters are user adjustable. There is also an online JavaScript version with access to data from most countries and states.

An initial value of R_0_=2.5 was taken from the March 20-21 case data for the US and within range of CDC and other estimates [1, 10]. We also conduct simulations at R_0_=3 to check sensitivity to this parameter.

For resource utilization the number of ventilators in the U.S. including reserves, alternatives (anesthesia machines) and older equipment is taken at 200,000 [10] and reduced to 100,000 as likely actually available. The number of ventilators in the world is harder to obtain. About 340,000 were identified at https://en.wikipedia.org/wiki/List_of_countries_by_hospital_beds but a number of large countries were listed as “unknown”. A rough estimate of 500,000 was assumed. A parameter for manufacture of additional ventilators accommodates announced intentions or running simulations to determine requirements.

The case ratio of total likely cases including undocumented ones to known reported has varied. Lower numbers are more critical due to the way the model calculates ventilator requirements. Higher numbers imply lower mortality than expected and achieve peaks more quickly. Published numbers began around 14% [11] which is a ratio of 7.1. Numbers eventually reached as high as 50 to 85 from randomized testing in Santa Clara County, California [12]. In New York State a range of 7 to 12 was evident from the Governor’s announcement on April 23 (see https://www.nytimes.com/2020/04/23/nyregion/coronavirus-new-york-update.html). Such testing is surprisingly slow to happen, but complicated by high rates of false positives. Yet case ratio is a critical parameter in estimating deaths [13]. While we simulate over the range 7 to 80, it would be useful to know what to expect. A value of 12 allows rough matching of deaths predicted by a Wharton model (viewed on May 12, 2020 at https://budgetmodel.wharton.upenn.edu/issues/2020/5/1/coronavirus-reopening-simulator), but while that is reasonable for the New York, New Jersey and Connecticut combined area, which we adopt as a principle sub-region NYNJCT, it does not fit the US data excluding that sub region. Many of our simulations are done at 15 with some lower and higher

A precise number for how long a case of COVID-19 lasts is not obtainable due to the wide variation. Data is complicated by regulatory requirements for waiting and testing. For matching public data on active cases 14 days is reasonable. But for matching known values of R_0_ and observed case growth rate, an average spreading period of 6 spreading days is used. Quite often R_0_ does not actually appear in SIR (Susceptible-Infected-Resistant) models, but instead the product of contact rate, infection rate, and disease period (or case duration). In order to have R_0_ directly appear in our model, we use R_0_ / (spreading-days) as the principle propagator coefficient.

The fraction of cases that require resources such as a ventilator is also important. We used 5.0% of known cases, or about half of critical cases, taken from Meng, et. al. [14]. Lower estimates and wide variety exist. Regionalization is also important as ventilators may not be distributed where needed. In another epidemic it may be some different resource. If no resource is critical, one must invent a “parameter” which is related to mortality to serve the model internals. In the case of COVID-19 we assume mortality can be estimated relative to the percent of patients on vents, and this is a regional parameter. The percentage may be greater than 100% since people not on vents may also die.

Mortality among known cases varies by city or region. The mortality rate is changing and likely to come down a great deal as it has been found patients respond better if just given oxygen not ventilation, various drug therapies are introduced, and there is a general learning curve. The current trend in US data is shown in Figure 2.

**Figure 2.**
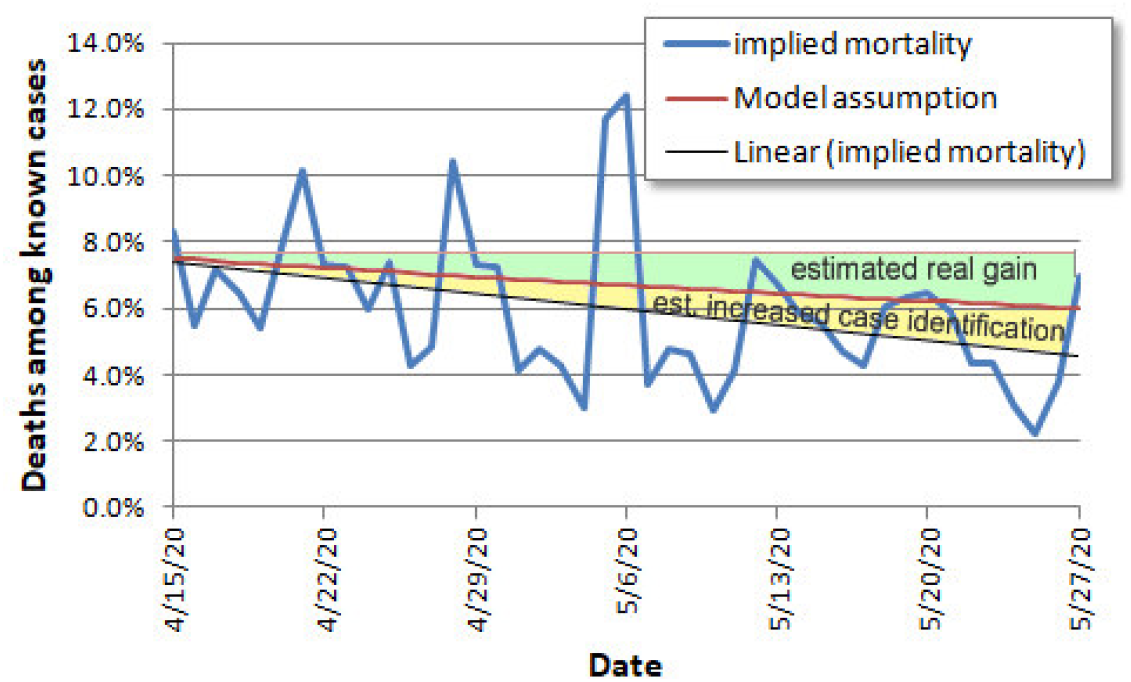
US COVID-19 mortality rate trend with linear fit, estimated from comparing daily reported deaths to new cases one week earlier. Model baseline assumption (red) is about half the gain, the rest attributed to increased case identification (estimate).

We assumed that improvement would taper exponentially amounting to 0.04% per day ending in September. This would produce a 75% improvement and a high advantage to putting cases off until September. Dividing by a case ratio of 15 that gives an initial total case mortality of 0.53% and a September estimated mortality of 0.13%. This compares with 0.1% often attributed to flu, though with vaccines fewer people may get flu.

Economic activity is reported as a function of the unlock schedule, and assumed to be proportional to degree of unlock easing (reopening). However, as distancing will be greater and personal protective equipment used (masks), the gain in economic activity per unit of contact will be greater than what was lost. We used an efficiency factor of 1.2 as an estimate of minimum expected gain. The parameter can easily be adjusted when using the model. For theoretical discussion, the number is not important as it does not affect infection behavior.

Five unlock dates are provided, each with its own percentage effectiveness.

Inputs are provided for a population annual growth factor and for the number of days immunity is expected to last. These are used for analyzing potential for recurrence or persistence using a 14 year simulation. Our simulations were run with 0.6% population growth for the US and 1.1% for the world, with a 730 day assumed immunity persistence. This has no noticeable effect on the short term simulation, only the recurrence check.

Inputs are provided for seasonality. These are *not* used for school or other cultural patterns which are more appropriately handled with planned unlock percentages. They are only for climate seasonality. Based on the previously mentioned 40% and 20% figures for New York and Florida, averaging them to 30% for the US, and reducing to 20% as an estimate of the amount not attributable to school, we arrive at a seasonality factor for the US. Only half the reduction is taken in the first and last months, which are assumed to be May and September. As 90% of the world population lives in the northern hemisphere, we used the same seasonality factor for the world.

### 2.2. Model dynamics

We use an SIR model engine [15] with parameter-driven forcing functions on R_0_. Spreading days converts R_0_ to a case propagation factor. During “lockdown” the reproductive factor is adjusted according to (a) the ratio of new cases from the previous day, (b) the herd immunity factor, and (c) the current seasonality factor. By this method a baseline R_0_ that reflects only social distancing and business closure is recorded. A moving average on this number prevents wild swings from anomalous data.

The use of the moving average is continued into the prediction phase where it represents gradual adjustment of the population to the new regime.

The model is designed to be instantiated with data prior to any unlock actions so a baseline 0% unlocked R_0_ can be established using the average for the previous week. In the case of COVID-19 there appeared to be a weekly pattern which might be some kind of data collection artifact of significant magnitude, and this averaging technique removes much of it.

When an unlock policy is established in the predictive model, the reproductive factor is biased toward the initial value and proportioned toward the last data-derived baseline reproductive factor according to the unlock effectiveness. Then it is reduced by the herd immunity factor. Calibrating unlock effectiveness, i.e. what policy will have what percentage unlock effect, is an important activity that is left to the user and local authorities and does not affect our theoretical investigation.

Each day actual data was used to replace predicted data. This affects the model’s integration base and the effective reproduction rate. The number of total cases, used for the herd immunity calculation, is calculated by the case ratio model parameter.

The effective reproductive factor for calculating the next day’s cases is either taken from the data, or for future projections calculated using the last empirical value while fully locked down, seasonality, percentage of susceptible individuals remaining, and the unlock percentage. Based on average case duration it updates the total active and resolved cases and calculates ventilator utilization.

## 3. Results

Figure 3 compares four world reopening schedules for COVID-19, also applicable to similar epidemics with no prior immunity in the population. The US is excluded and will be addressed separately.

**Figure 3.**
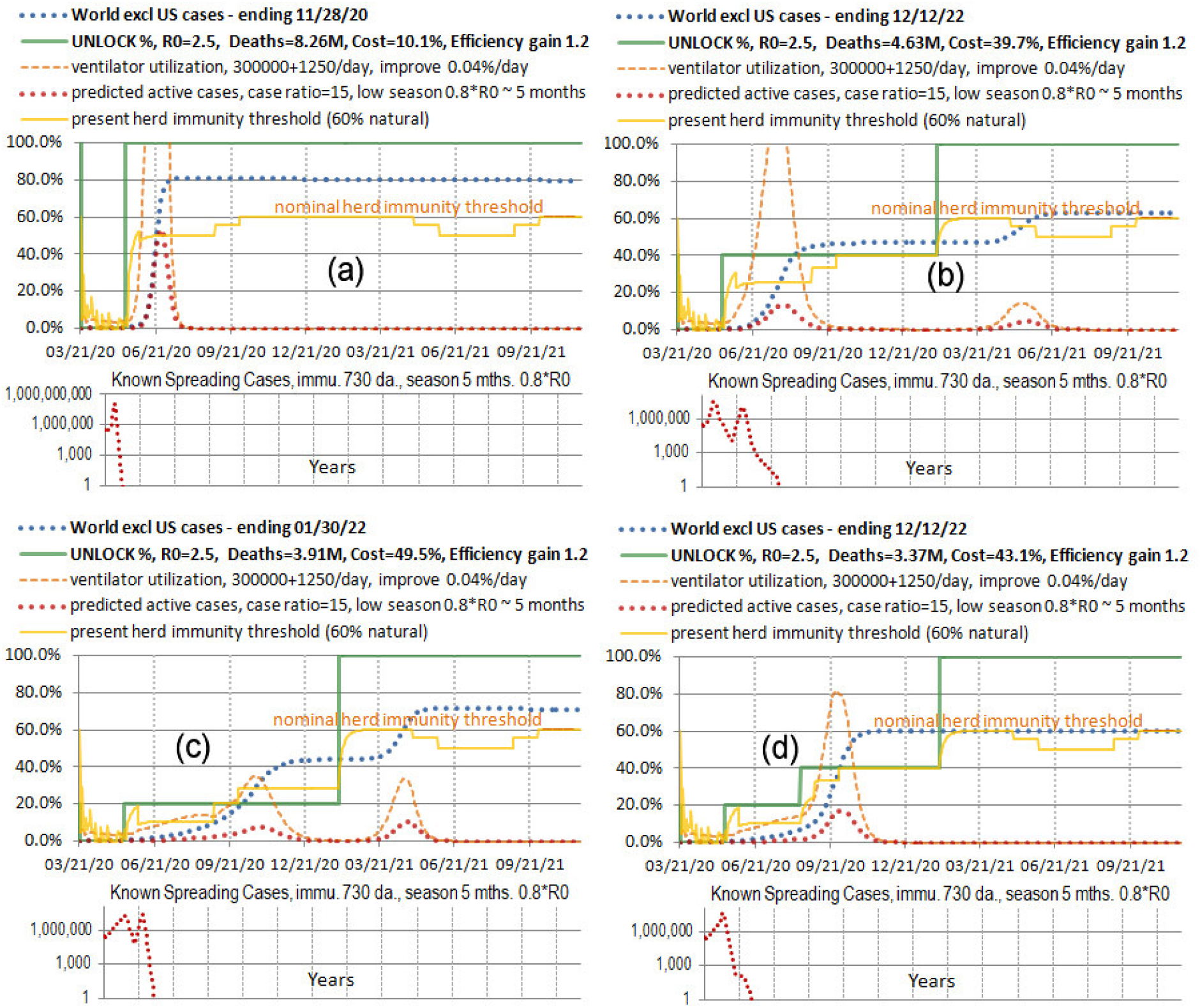
COVID-19 World reopening schedules for R0=2.5 and case ratio=15 (a) immediate full unlock, (b) 40% immediate and 100% in February 2021, (c) 20% immediate and 100% in February, (d) 20-40% multi-step unlock and 100% in February.

For replication factor R_0_ up to about 4 we see similar results. For R_0_=4 nearly herd immunity is reached if there is even a two week period before lockdown. All higher replication rates over-shoot herd immunity if there is a two week window, and replication rates over 7 infect 99% of the population. This assumes homogeneous contact, a characteristic of SIR models, a sort of worst spreading case. Spreading may take longer if it has to reach isolated populations are travel through a geographically constrained region. Equal spreading in all world climates is also assumed. So there are a lot of caveats. It is a general guide.

The deaths and economic cost for the above cases including alternative R_0_ and case ratio values are in Table 1 for these values and for alternate values of R_0_ and case ratio.

**Table 1.** SIR-projected world deaths and economic impact for four reopening schedules of Figure 3 Baseline R_0_=2.5 and case ratio 15. Alternatives in right two columns.

| Schedule | Deaths<br>*in millions* | Economic<br>Impact<br>*as % of 18*<br>*month lockdown* | Deaths for<br>$R_0=3$ | Deaths for<br>case ratio = 50 |
| --- | --- | --- | --- | --- |
| (a) immediate | 8.26M | 10% | 9.57M | 2.58M |
| (b) 40% - 100% | 4.63M | 40% | 6.09M* | 1.56M |
| (c) 20% - 100% | 3.91M | 50% | 4.82M | 1.35M |
| (d) 20-40-100% | 3.37M | 43% | 4.19M | 1.09M |

\* Indicates ventilator capacity may be exceeded

The economic impact is only a function of the unlock degree and schedule. Generally case ratio does not change the trajectory of cases, but if cases are unknown because they are mild then it dilutes both mortality and resource utilization proportionately (e.g. double case ratio and half mortality). Case ratio does not remain constant. The table is for some value expected when the simulation is run. A higher replication factor will create more cases and if they have the same mortality (not always true) will result in more deaths, but only slightly more. It is not proportional.

Notice that the total cases (blue dotted line) are about the same in (b) and (d), but the deaths are higher in (b) because they are earlier, where we have assumed higher mortality. The immunity threshold is drawn (yellow) to show the theoretical stopping point for total cases at each level of lockdown, based on the value of R_0_ associated with that level. It is apparent that overshoot is a problem even for modest amounts of reopening such as the 20% case in (c).

The lowest deaths in (d) is from a combination of elimination of overshoot and postponment of most cases until September when mortality is presumed to be lower. August 15 unlock was chosen to support school. The difference between comparable schedules (with cases in the September time frame) of overshoot for 20% then 100% unlock (c) and the best targeting of population immunity (d) is 0.54 million deaths avoided.

If we remain fully locked down until an August 15 100% unlock (not in the figure, date chosen to support school in the fall) the overshoot will be to the 82% level and 4.77 million deaths, with 1.44 million avoided by using a partial unlock approach with overshoot avoidance. The reason an overshoot in the fall is higher than an overshoot immediately is because we are already into the low season for the virus in May, and reducing R_0_ by the seasonal factor. In the fall it is full strength. Option (c) is both more costly, more lethal, and does not provide additional easing to allow a fall school semester. A 10% version of (c) still sickens 40% of the population by March 2021.

If the anticipated mortality reduction is more modest, a quarter of that projected here, the worst case deaths at a case ratio of 15 becomes 14.2 million, and overshoot reduction of schedule (d) reduces that to 9.1 million, a 36% reduction. For case ratio of 7, which was initially supposed, the numbers would be 30 million with full overshoot and 20 million with schedule (d).

### 3.1. Overshoot Analysis

Handel et. al. [9] say that to avoid overshoot, cases should vanish as one approaches the herd immunity threshold. This assumes we know what that threshold is, and have very fine control over cases. They suggest “adaptive” control. Overshoot arises in the following way. At the immunity threshold, effective R_0_ is 1.0. If 1% of a population is sick, they will still sicken another 1% and you have overshoot 1%. Then R_0_ is reduced a little more but some additional people get sick, and so forth.

Figure 4 (a,b,c) gives an idea of how much overshoot to expect from a 10% single unlock step for different values of natural R_0_ in an epidemic with case data similar to COVID-19. The bottom row (d,e,f) compares 20% and 40% unlock steps, then in (f) adds 20% seasonality. The partial unlock population immunity value (yellow) is low in the early part of the graph because it is from actual data, which hasn’t yet responded.

**Figure 4.**
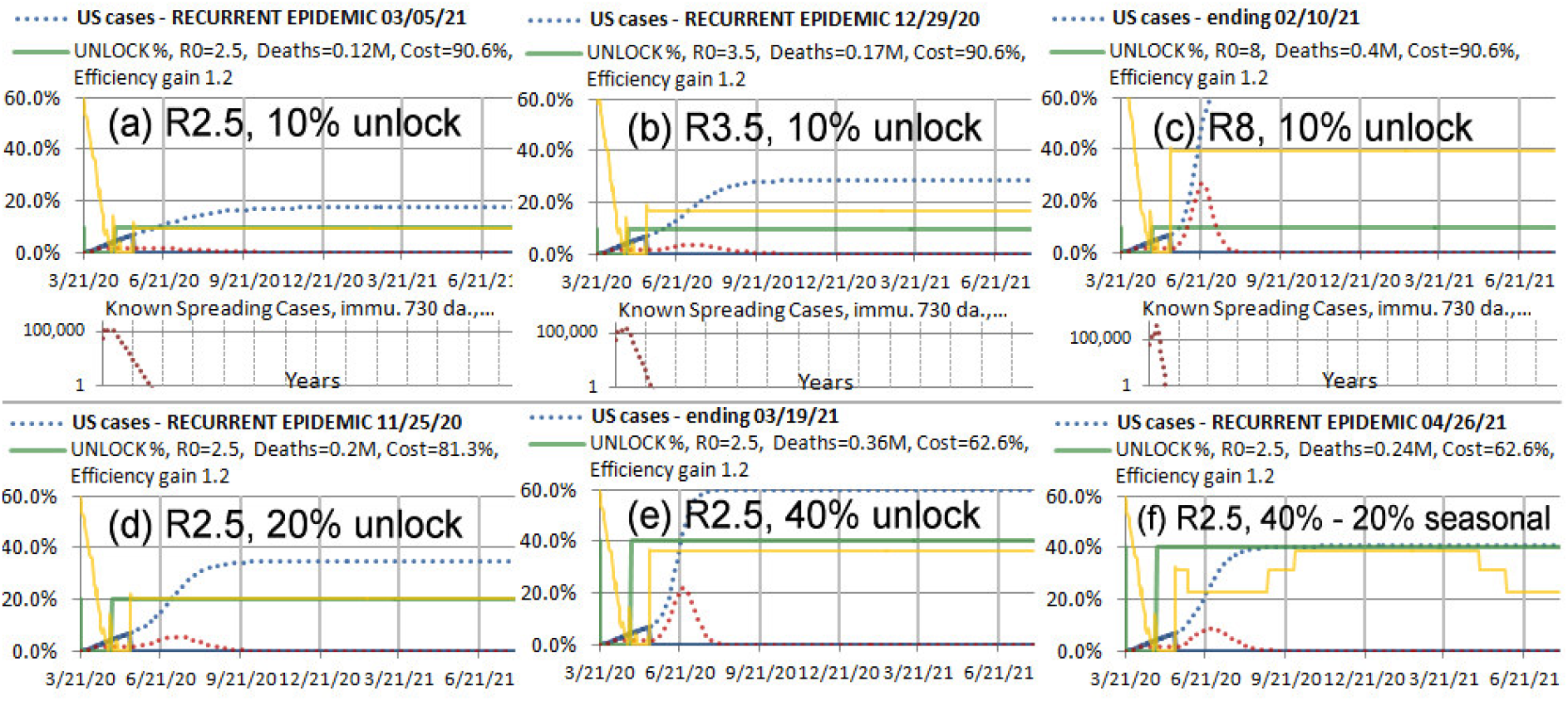
Expected overshoot from COVID-19 cases through 5/17 (a), (b), (c) R_0_=2.5 (likely), 3.5 and 8 (some other pathogen) for 10% unlock, no seasonality (d), (e) 20% and 40% unlock for R_0_=2.5, no seasonality (f) 40% unlock for R_0_=2.5, with 20% seasonality

Seasonality is ommitted from most of the figures because it makes an effective two step unlock and depending on value and timing, the overshoot may disapper or be exaggerated. In (f) the seasonality for the actual May 15 COVID-19 unlock in the US is depicted, but 40% is not necessarily a projection of what this unlock will be. It might be more or less. There is a mix of unlock strategies in the US, but states that do unlock are having a hard time controlling the degree because as a practical matter people are not able to work unless childcare is unlocked, and children in the US are not at this time disciplined to wear face masks.

For R_0_=2.5 and low initial case numbers, it appears an overshoot from a single step unlock is about 60% above the population immunity threshold for that level of unlock. It is at least twice that for R_0_=3.5, and for R_0_=8 a 10% unlock fully unleashes the pathogen. Since the empirical calibration of lockdown is based on COVID-19 data, it is not clear current measures would have been effective against a high R_0_ pathogen.

Note that for low to moderate unlock levels at R_0_=2.5, they are approximately equal to immunity thresholds. This appears to be happenstance as it does not hold for other R_0_ values. It is convenient for thinking quickly about COVID-19 but can’t be counted on for other pathogens.

### 3.2. US analysis

Even the US is not managed as a single region. As of late May 2020, 58% of the country was on strict lockdown, while 17% have resumed pre-coronavirus activities in a nationwide poll (see https://nypost.com/2020/05/15/social-distancing-on-the-decline-in-the-us-poll/). Some describe themselves as only mostly isolating. It is partly on this basis that the authors have been assuming a 20% level of unlock for May 15. Simulations for the NYNJCT sub-region, with a greater number of people already immune, show that cases will decline for any unlock of 25% or below. For the rest of the US a 20% unlock will produce only a slight rise in cases. This is mostly due to the assumed 20% percent seasonal factor, not counting schools which were already closed in March.

The overshoot situation for 20% differs between regions. In NYNJCT a high number of cases has them near enough to the 20% immunity threshold that seasonal transition in the fall provides an adequate two step unlock to avoid overshoot. In the rest of the US a rebound would occur even if unlock were maintained at 20% through the fall. However the situation is sensitive to levels, with different outcomes at 10% or 30%, and sensitive to the seasonality factor, for which we only have a rough estimate and which varies by region. The situation is not favorable for managing an epidemic with a single or double step unlock. Figure 5 (a) shows a 20% unlock for the entire US followed by an additional 15% in August for school and other fall activities.

**Figure 5.**
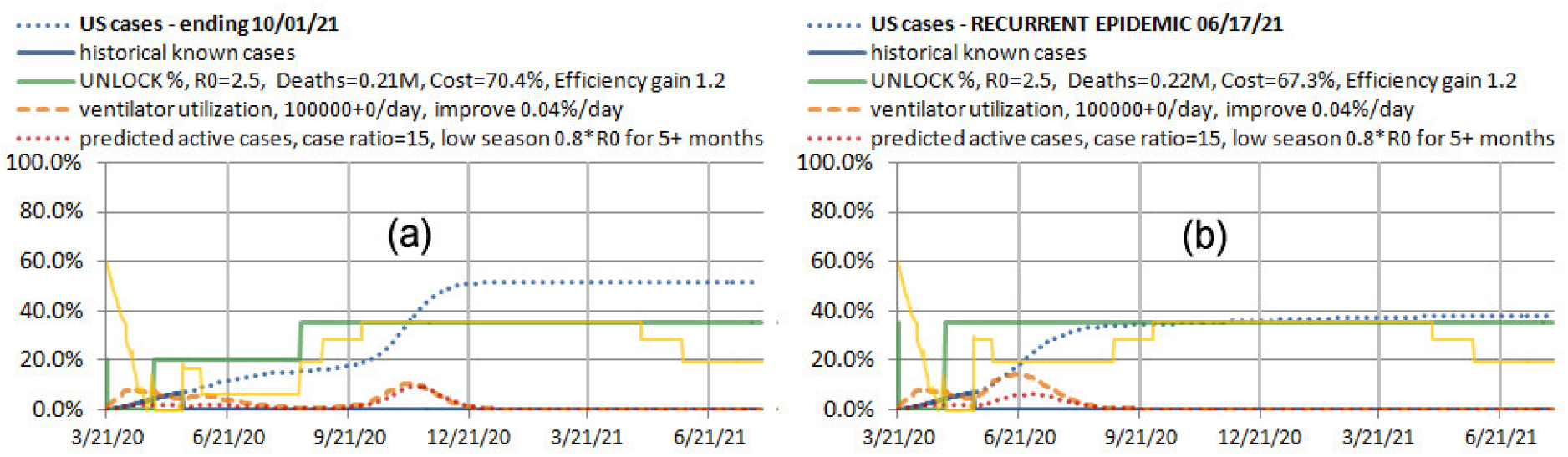
Modest unlock schedules for the US, (a) 20% then 35% for school, (b) 35% now.

The overshoot in 5(a) is so bad the country may as well go for natural herd immunity. By unlocking 35% immediately in Figure 5(b) the seasonality provides about the right sized first step so that effectively we have a two step unlock which targets the population immunity threshold for 35%. Deaths occur earlier at the higher mortality rate, but overshoot avoidance is so efficient that the total deaths are not appreciably different. However the costs of both are high. There is insufficient relief. The final straw is that these strategies are sensitive to variation in seasonality, timing, R_0_, case ratio, and unlock percentage. Just the difference between (a) and (b) conveys an idea how much change a rather modest amount of unlock can make in a two step schedule.

As an aside, it appears to the authors that maintaining a full lockdown to await a vaccine is not going to be accepted in the US and several other countries. It results in low cases, reduced fear, and people have already decided to venture out even if their state governments have not relented yet. Due to the summer low season, there will not be enough rise in cases to deter them. This is a reality the authors feel those trying to manage an epidemic must accept if they are to be effective in helping people. In a country like the US it is not the time or place for ideology and for making choices for people. High control strategies may work for some countries.

So what is a realistic strategy for the US, one that isn’t too prone to failure if some parameter varies a little? A practical strategy is to use a multi-step unlock which more closely guides the trajectory of total cases, washing out the effects of overshoot of intermediate levels. A sensitivity analysis of this is shown in Figure 6 over a range of parameters.

**Figure 6.**
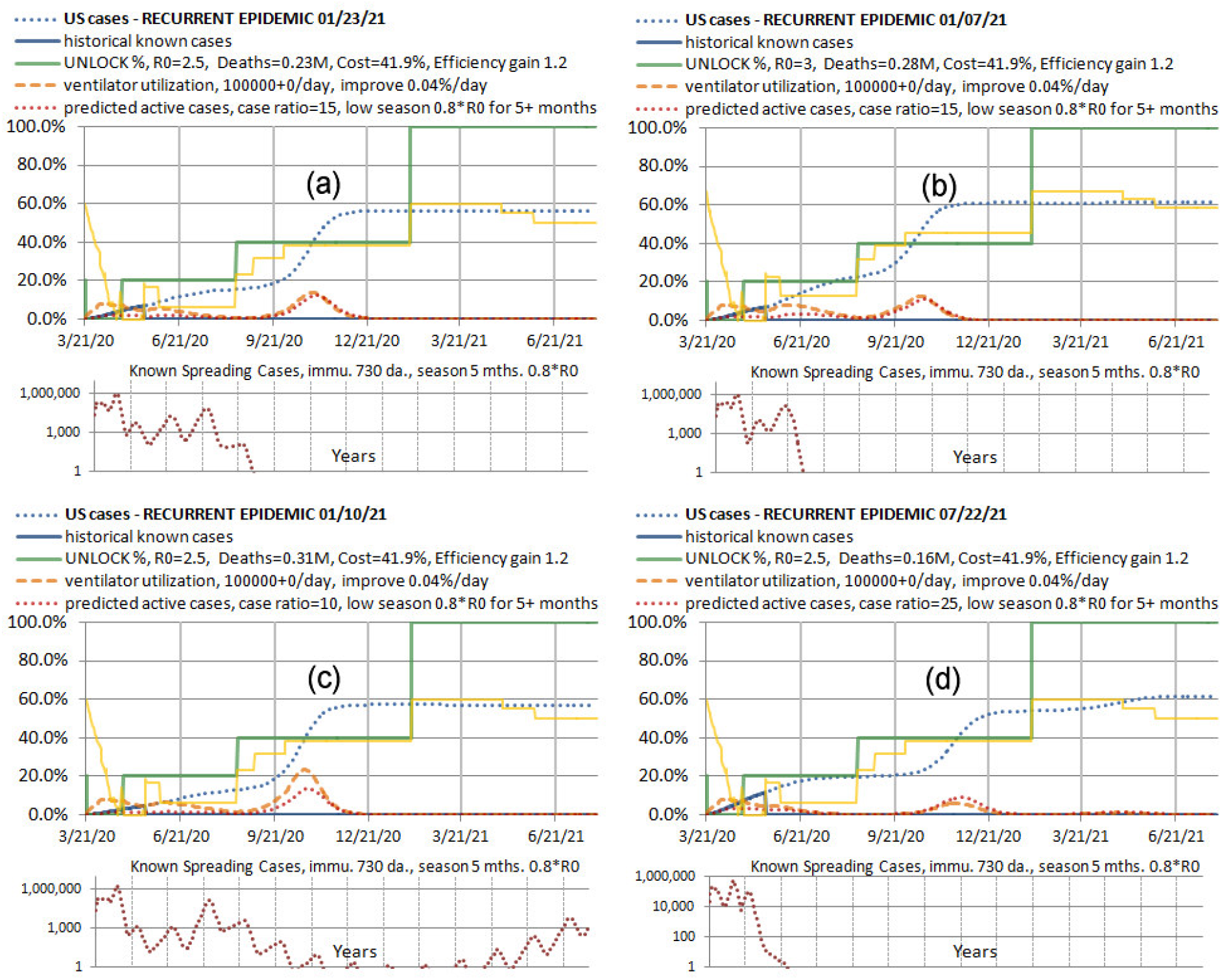
Sensitivity analysis for a three step US unlock targeting natural immunity. (a) Baseline case with R_0_=2.5 and case ration 15. (b) Increase R_0_ to 3. (c) Low case ratio of 10. (d) High case ratio of 25.

All of the cases in Figure 6 perform as they should. This seems remarkable to the authors after two months of pushing on one part of the cases trajectory only to have a rebound pop out somewhere else. Changes in timing of unlocks are not shown but those simulations were done and it is not particularly sensitive to a month one way or another. If the case ratio is over 25, there will be increasing overshoot. However these scenarios have much lower mortality in general, so if it has to fail somewhere that is the least damaging place. The schedule is designed to work with seasonality. If seasonality is higher than we’ve assumed, it works a little better, and vice versa. If seasonality is zero, it overshoots to the 70% level. Furthermore the economic costs are barely more than half the conservative cases of Figure 6, and the death totals for comparable scenarios (case ratio of 15) are very close, a small price to pay for robustness to variations variations that might cause greater deaths.

## 4. Results after Two Months (mid-July 2020)

The preceding analysis was based on data and information available on or before May 19, 2020. The preliminary results were made available but formal review was postponed for two months to see how actual events would play out, and thus better assess the validity of the proposed methods. It is a fact that an epidemic moves at its own speed. It does not stop and allow for clinical trials, peer review, or any other of the niceties of science. However, the situation two months later is somewhat similar due to the success of lockdown policies at holding infections, if not in stasis, at least at levels significantly below population immunity. Thus the potential for overshoot and unnecessary deaths, as we will see, still exists. Despite testing, case ratios are still as uncertain as before. There just isn’t emphasis on the kind of random sample testing that would settle case ratio. Without case ratio, modeling is guesswork. This is a deficiency that should be corrected both now and in the beginning of future pandemics or epidemics. The significant thing that has changed in the intervening two months is that two months of regional data for all kinds of countries and regions during easing of lockdown is available, making possible a resonable calibration of R_0_ and unlock percentages vis a vis what populations and governments are likely to actually do. Figure 7 shows four US states.

**Figure 7.**
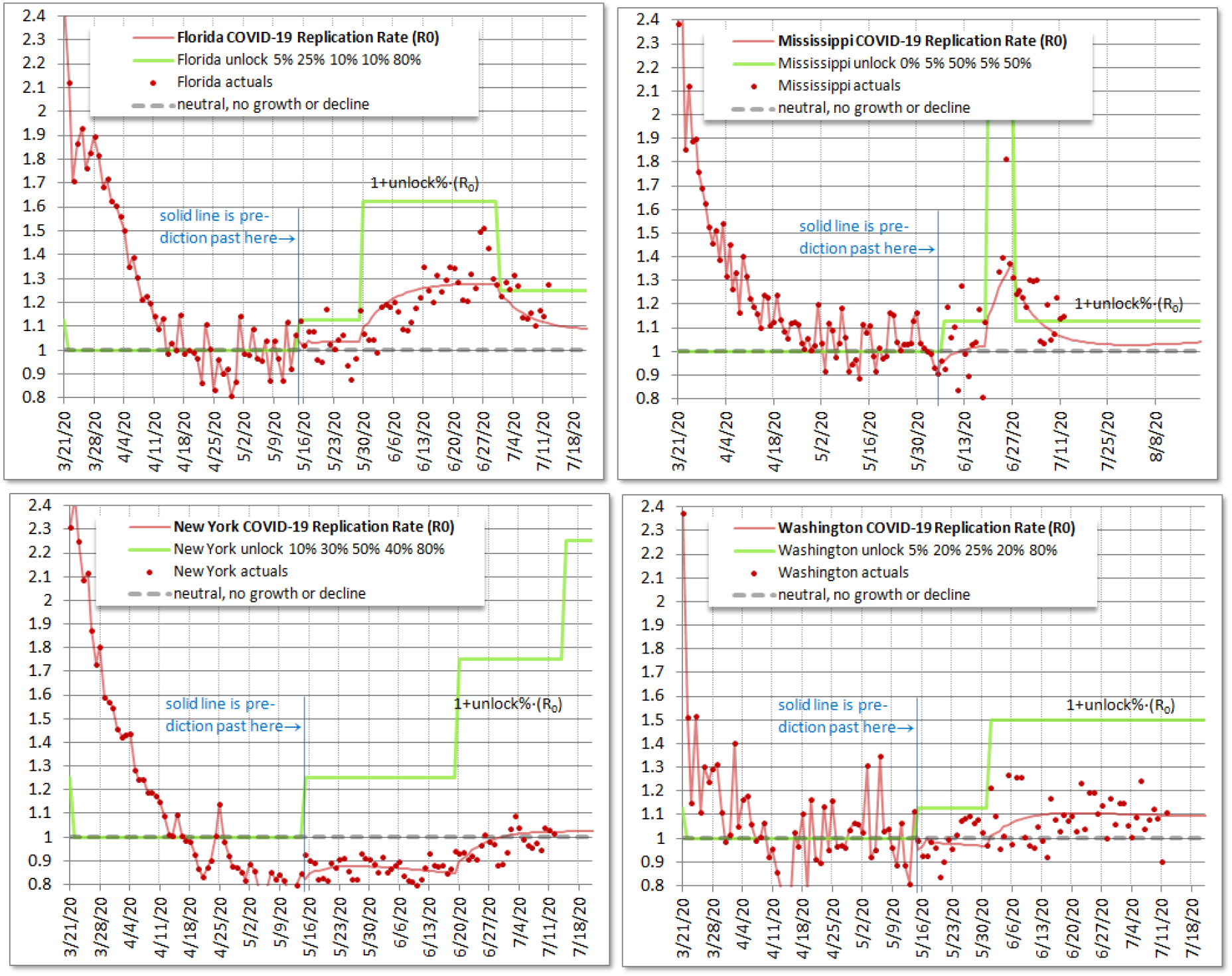
Daily R_0_ for four representative US states as of July 13, 2020, with a model forecast R_0_ (solid line) from May 15 data (approximate lowest R_0_) based on assumed unlock levels (for calibration of unlock levels).

**Figure 8.**
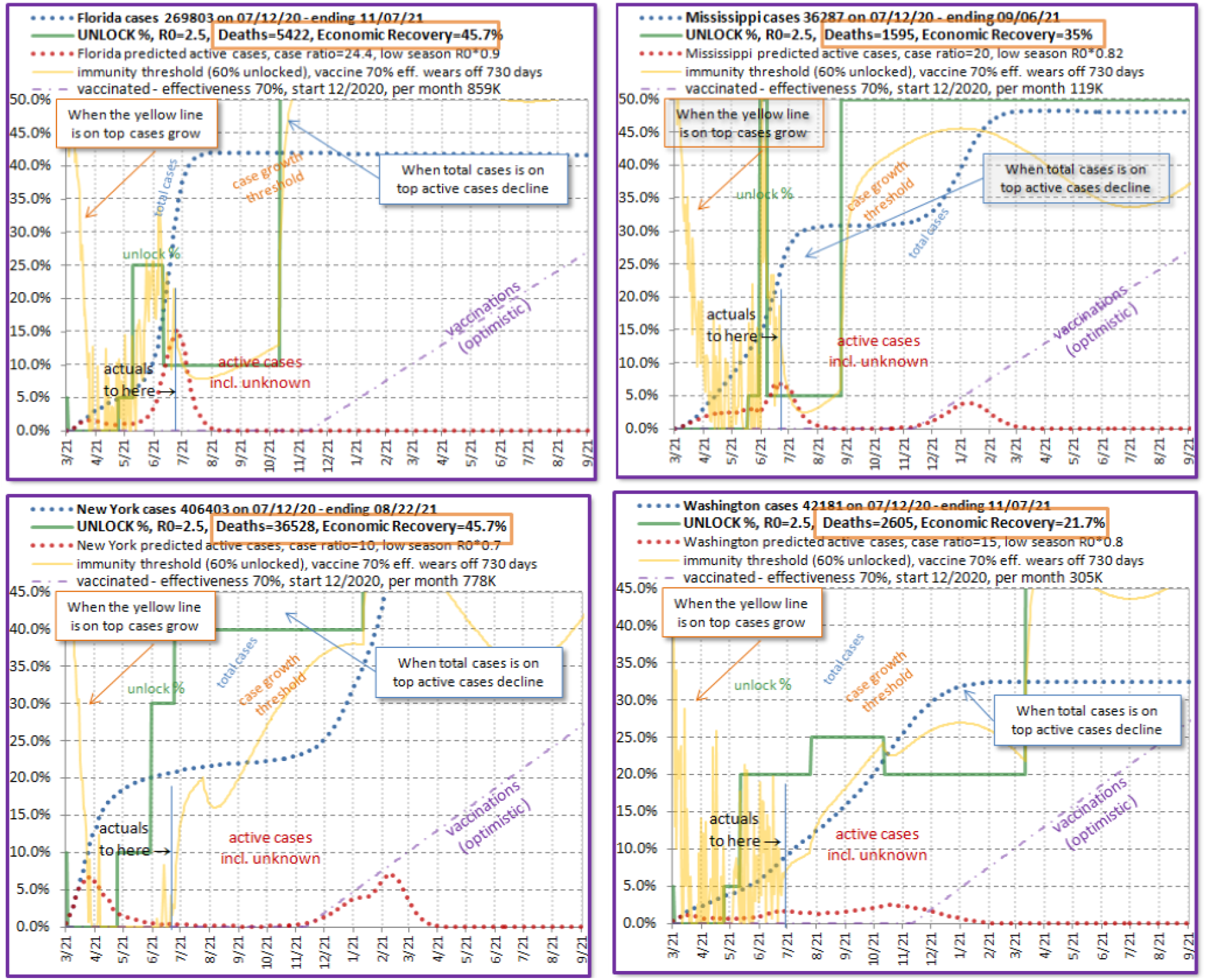
Comprehensive forecasts for the four US states of Figure 7. FLORIDA (upper left) 45% economic recovery, relock, no rebound, no overshoot MISSISSIPPI (upper right) 35% recovery, relock, post-vaccine rebound, overshoot. NEW YORK (lower left) 45% recovery, no relock, post-vaccine rebound, overshoot. WASHINGTON (lower right) 21% recovery, mild relock, no rebound, some overshoot.

Two states show evidence of re-lock and two don’t despite announcements by the states of re-lock. Many other states and countries are typical of these patterns.

Each chart shows at least two levels of unlock compared with actual data, with three levels in the case of the top two which have a re-lock. The method of calibrating the unlock levels was as follows. Using a macro, the model was run with data through only May 15th, which for most US states included the lowest R_0_ and allowed the model to calibrate its fully locked R_0_ value. This resulted in a prediction based on assumed unlock levels after May 15th, the solid line which does not specifically connect daily points.

The unlock levels were then adjusted so that the solid line ran approximately through the middle of the points. The uncertainties in unlock and other parameters are so large it was not deemed advantageous to use some kind of mathematical curve fit, just an optical centering. Generally this allows unlock levels to be determined within 5%, closer than we expected. The full data are then restored for future modeling, and these charts are saved for reference demonstration that the unlock levels between May 15 and July 13 are valid representations of what was going on in the regions during that period. Future unlock levels can be meaningfully estimated based on those. For example, some additional unlock for school, perhaps half as much again as the general social unlock, might be added in mid-August.

Or if there is a large case peak forecast, enough re-lock to contain it will be forecast. This estimation is made manually on the basis of risk homeostasis, i.e. that the society has become accustomed to a certain level of cases, and thus risk of social and business interaction, but will become alarmed if cases rapidly increase. This is in fact what has been stated by officials and media as explanations for re-lock, so we apply the technique in forecasting.

Examples of good and bad economic recovery, without and without overshoot, with and without re-lock are evident in nearly every combination. For details, see the figure caption. For Florida, a high case ratio was used based in published test positivity. But it is still a guess as tests were not random. This is the reason Florida experiences no rebound whatever, though it would likely be mild in any case as Florida is intent on letting cases build and eventually will get to herd immunity regardless of the case ratio. The question is how fast and will it overshoot? We think no overshoots will occur during the summer in the US because the public becomes alarmed and officials relock. During the summer, with the help of seasonality, it is easier to relock. States that have already had a lot of cases get a slight boost from the amount of immunity they have accumulated. If 5% of the population has or has-had COVID-19, then there is an additional 5% unlock possible while obtaining the same R_0_ value. In Florida it is practically summer all year, and humid, so it is easy to relock even in December or January. Florida likely will avoid overshoot.

For the other three states, we predict some degree of overshoot. For Washington, we think the relock will be just enough to keep cases from growing. The maintenance of too high a level of cases as population immunity is approached will cause a mild overshoot. This, the reader should thoroughly understand, is mainly an estimate of human behavior and peculiar to the state of Washington (though somewhat prevalent on the west coast of the US). It is not a constraint of the SIR model or climate in Washington. There is plenty of room for them to relock. However, they already have the lowest economic recovery of the four states presented by a large margin, and there will be some public tension between case control and economic recovery.

Our estimates for when vaccination will start and its effectiveness are overly generous in order that our modeling may not be criticized as predicting deaths and economic loss because it is too stingy with the vaccine. It is at least possible large scale trials could begin in December, and possibly vaccination of medical personnel. Rather than the 70% effectiveness we have assumed, which is roughly equivalent to the advantage of mask wearing, persons who have had access to early trial data (which is not published at this point) estimate the effectiveness more likely in the range of 40-60% (https://www.newsweek.com/first-covid-19-vaccine-flu-shot-1516343). The forecasts are based on the idea that behaviorally, people will want to try unlocking to a greater degree when vaccination starts, or after it has been going a few months. But at first only a few people have been vaccinated with a weakly effective vaccine, and this results in significant rebounds for two of the states, with overshoot associated with each.

## 5. Discussion

The three (or more) step approach seems to be more stable because it guides the total cases using the natural population immunity associated with each level of unlock.

While virus evolution is not a certain science, there is reason to believe viruses are rapidly selected for increased transmission (R_0_) [16]. A long delay in building immunity to the virus and driving it mostly out of the human population leaves a large number of active cases present in which the virus is being selected to overcome the near-1.0 R_0_ condition of social distancing, eventually making the virus more difficult to control by this method. A long period of persistence or multiple recurrences, though involving a smaller number of cases, also provides the opportunity for virus evolution.

The way in which seasonality provides a natural two-step modulation of overshoot is interesting and bears further investigation. This may assist pathogens in obtaining a near undershoot condition and establishing themselves as recurrent or even endemic. While the authors didn’t focus on this issue in this paper, we have many simulations showing recurrences oscillating forever. Human targeting of the threshold, which is imprecisely known, makes such recurrences more likely. Some epidemiologists feel that the virus will hide somewhere anyway. Certainly the world is not homogeneous. However, reintroductions may be easier to contain than diffuse recurrences in multiple locations. There are few vaccines that have been distributed effectively throughout the world, and none in a short time. Over-doing immunity by vaccine or naturally seems a reasonable goal.

The authors urge government officials to be utterly straightforward about the goals and likely length of lockdowns, and to consider freezing financial obligations and employment status while such measures are in place. We do not expect people to be fired or loans foreclosed because of other civil emergency response actions.

It has been suggested that COVID-19 and its related cousins (SARS, MERS) do not kill directly, as the virus count has already passed the peak when death occurs, but the *coup de grâce* is delivered by the immune system fighting back too hard [17, 18]. Is that how our society and civilization is going to die, by fighting too hard?

A list of preparations for epidemic response provided by Smith and Fraser [19] includes “*data and analytics capabilities; maintain and expand our state-of-the-art public health laboratory capacity; continue building a workforce of trusted, expert, public health professionals; sustain our capacity to rapidly respond to outbreaks at their source; and assure a strong global and domestic preparedness capacity*”. Are the necessary actions to lock down economies without destroying them, make difficult and practical tradeoffs on behalf of the public, and implement phased reopening on the list?

Complicated things like stopping and starting an economy do not happen without rehearsal, involving both the officials who must coordinate response, and ordinary citizens. But matters outside the purview of epidemiology and in the realm of massive government action are not addressed.

Each discipline is thinking only within its boundaries, yet giving advice across those boundaries that has profound effects and global reach

### Planning is not nearly sufficient

Ordinary disasters like storms and fires require *rehearsal*. Military operations require *war games*. Already in direct government outlays the US has spent more than double the cost of the Iraq war, longest in its history, on COVID-19 response. Preparation for future disasters of such high probability requires no less attention.

It has been suggested that if there is no cure or vaccine for an epidemic or pandemic the best possible option may be social distancing and isolation [20]. However Figure 4 and 5 along with worldwide public reaction after two months of lockdown make it clear that to simply specify social distancing will not minimize deaths. As the need for longer periods of isolation become apparent and the necessity of providing food (even opening COVID-infected meatpacking plants) and paying rent (not every country can provide a multi-trillion dollar relief package) rise to the fore, some degree of reopening is inevitable. It is not an adequate response for public health officials to wash their hands of the matter, their advice not followed. Effective advice on how to do what is required, and achieve a practical outcome, perhaps an outcome dictated by political processes rather than epidemiological considerations, is required from the public health community.

## 6. Conclusion

We have shown that overshoot, often aggravated by sensitivity to unknown parameters, can dominate the mortality rates of an epidemic, ruining otherwise attractive strategies. We showed that two step unlock is effective at minimizing overshoot, but too sensitive to unknown variables. Three or more steps provide more robustness to parameter variation by more closely guiding the total cases trajectory with gradually increasing population immunity. Seasonality must be considered in planning the steps.

This paper quantifies mortality costs associated with following a less than optimal unlock strategy and identifies likely case creep even in the most conservative strategy. We provide specific tools and theoretical understanding for finding and following a near-optimal strategy, whatever goal society adopts.

And finally, we showed that well into the unlock stage, the problem of overshoot persists, and likely will persist even after vaccination begins.

## Data Availability

The spreadsheet model with data to replicate simulations in the paper will be uploaded with the paper

http://shulerresearch.org/covid19.htm

